## Supplementary_Material for "App-based COVID-19 syndromic surveillance and prediction of hospital admissions: The COVID Symptom Study Sweden"

**Question updates in the COVID Symptom Study Sweden baseline survey**

Upon downloading the COVID Symptom Study Sweden (CSSS) app and providing informed consent, participants were asked to complete a health survey including pre-existing health conditions. The baseline health survey included questions on diabetes and lung disease that were later expanded to include disease subtypes in late June 2020. Prior to that, participants could only respond yes/no to presence of “diabetes” and “lung disease”. Following the updates, participants who had previously indicated presence of diabetes and/or lung disease could specify type of disease, and new participants were asked the more detailed questions upon registration. Consequently, participants who did not submit a single daily report after June 24, 2020 were all labelled as having “type not specified” diabetes and/or lung disease. Other baseline questions, for example on dietary supplements or physical distancing, were introduced in June 2020 but subsequently removed from the app in September/October 2020. Information on all questionnaire variables both from the baseline health survey and the daily report, including information on whether the question was available at launch, or the date when the question was added and/or removed, are presented in Supplementary Table 6.

**National population and health registers**

In order to compare CSSS estimates with national health register disease data, we extracted an anonymised population-based dataset from official national population and health registers held by Statistics Sweden and the Swedish Board of Health and Welfare, outlined below. Linkages between national registers were enabled by personal identification numbers, unique 12-digit numbers assigned to all individuals in Sweden at birth or immigration. We extracted information on sex, year of birth, and home address postal code from the Total Population Register held by Statistics Sweden, and combined this information with data from the three health registers outlined below. The national register data were not linked to study participants in COVID Symptom Study Sweden.

**SmiNet**

The Swedish government classified COVID-19 as a communicable disease dangerous to society as well as to public health on February 1, 2020[1]. In accordance with the Communicable Diseases Act (2004:168) and the Communicable Diseases Ordinance (2004:255), it thereby became mandatory for all clinical laboratories in Sweden to report positive PCR tests for SARS-CoV-2 to SmiNet, an electronic notification system of communicable diseases maintained by the Public Health Agency of Sweden. We obtained data on all individuals ≥18 years with a positive test registered in SmiNet from January 1, 2020 to January 4, 2021. The first date variable, date of testing, was missing in 12.5% of all positive COVID-19 tests. The majority of those tests however had a valid date of arrival, the second date variable at the clinical laboratory where they were subsequently analysed. The third date variable, date of registration in SmiNet, was available for all tests. "Date of test" was thus imputed using the other two date variables, using multiple imputation by chained equations, then extracting the first imputation. The imputation model used was a linear regression, with 'date of arrival' and 'date of registration' modelled as restricted cubic splines with four knots each, placed on the 5th, 35th, 65th and 95th percentiles. The imputed times were rounded to the nearest date.

**The National Patient Register**

The National Patient Register, maintained by the National Board of Health and Welfare in Sweden, collects information on patient diagnoses from inpatient care as well as from outpatient visits to specialist care. Diagnoses are classified according to the International Statistical Classification of Diseases and Related Health Problems Tenth Revision (ICD-10). Swedish healthcare utilizes the two emergency ICD-10 codes for COVID-19 U07.1 (COVID-19, confirmed by laboratory testing) and U07.2 (COVID-19, virus not identified), the latter assigned to a clinical or epidemiological diagnosis of

COVID-19 where laboratory confirmation is inconclusive or not available. We extracted information for all individuals ≥18 years hospitalized with a first diagnosis of U07.1 or U07.2 from January 1, 2020 to January 4, 2021.

**Socioeconomic variables**

We obtained information on an aggregate level for all postal code areas in Sweden from Statistics Sweden for individuals ≥18 years. Data included sex distribution, population size, highest achieved education level in the population 25-64 years (categorized as compulsory/primary education only, secondary education, or university education), proportion engaged in work or studies in the population 20-64 years (categorized as occupied with work and/or studies, or not occupied with either work or studies), median yearly net income for the population ≥20 years (in 10,000 SEK), and proportion of population ≥18 years with foreign background. Foreign background was defined as born in a country other than Sweden, and/or born in Sweden but with both parents born in another country than Sweden.

We further calculated population density for all postal code areas in Sweden as the total number of inhabitants per square kilometres. The surface area of each postal code was estimated using shapefiles originating from an external company, Postnummerservice Norden AB, which contained detailed information on five-digit postal code area boundaries.

**National COVID-19 point prevalence surveys**

The Swedish Public Health Agency conducted five national surveys of point prevalence of COVID-19 in 2020, in April, May, August, September and December. We utilized results from the survey in May 2020 which was the first after the launch of COVID Symptom Study Sweden[2]. That survey was performed in the previously established population-based national cohort The Health Report ("Hälsorapporten") and included children as well as adults (n=2,957). Participants were sent self-test sampling kits, instructions for swabs, and also a symptom questionnaire. For children, their legal guardians were recommended to perform the swabs. Samples were collected by members of the

Swedish Armed Forces, and analysed at medical laboratories using PCR. The results were weighted for the selection of participants into The Health Report, the participation in the sampling effort, and the age and sex distribution of the Swedish population.

**NOVUS**

NOVUS conducts surveys using the NOVUS Sweden Panel, which includes approximately 45,000 panellists randomly recruited to be representative of Sweden in terms of age, sex, and region of residence. Recruitment is mainly done through randomly selected telephone interviews, in combination with targeted personal invitations to under-represented target groups. Personal invitations are sent by postal mail to individuals without a registered phone number, and text messages are sent to individuals who do not answer their phone. Self-recruitment is not possible. Panellists who have not responded to at least one survey during the last three months are replaced. When performing surveys, NOVUS invites a representative third of the NOVUS Sweden Panel to participate.

When we compared the symptom prevalence reported in CSSS to the symptom prevalence reported in the NOVUS dataset was split in two parts, one from March 23 to December 8, 2020 which consisted of 32,232 individuals (50·5% women, median age of 53 years (IQR 39-66) and the second from December 9, 2020 to February 10, 2021 which consisted of 8,630 individuals (50% women, median age of 53 years (IQR 39-67)).

**CRUSH Covid**

CRUSH Covid is currently an ongoing (as of October 2021) interdisciplinary research project on COVID-19 in the healthcare region of Uppsala, conducted by Region Uppsala in collaboration with Uppsala University (more information available at uu.se/crushcovid). From October 18, 2020, all individuals (≥18 years) who do a PCR-test of COVID-19 at any of the testing stations in Region Uppsala can voluntarily participate in the CRUSH Covid Survey. Out of the 2,116 participants who had participated in the CRUSH Covid survey until February 10, 2021, 943 individuals completed the survey and the COVID-19 test on the same day, reported at least one of the symptoms included in the CSSS model training, and had a conclusive test result. Of those 943 individuals, 67% were women (n=631), the overall median age was 41 years (IQR 30-51), and 15·3% (n=144) had a positive PCR test.

### METHODS

#### Neighbourhood Deprivation Index

We calculated a neighbourhood deprivation index (NDI) for each region as well as each postal code area in Sweden, by combining regional or postal code area level information on the proportion of adult inhabitants occupied with work or studies, the proportion with university education, and the proportion of adult individuals with an income in the lowest quartile of the nationwide distribution. A principal component analysis, based on the correlation matrix and weighted by the adult population in each region or postal code area, was conducted on these three variables. The first principal component was normalized by dividing the component by the square root of its eigenvalue. The interpretation of NDI is thus that the lower the NDI, the more disadvantageous the overall socioeconomic position for that area.

#### The nested tenfold cross-validation of the model developed in Step 1 to estimate individual probability of COVID-19

In both the main model and the time-dependent model, we performed internal evaluation of discrimination and calibration by applying nested ten-fold cross-validation within the dataset from April 29 to December 31, 2020. In a nested ten-fold cross-validation, Oppedal et al[3] outlined an outer and an inner loop of iterations, see Figure 5 in their paper. For each of ten iterations in the outer loop, we ran a LASSO on nine folds constituting the ”training data” and generated symptomatic COVID-19-probabilities for the individuals in the test data (the last fold). The shrinkage parameter for each LASSO model was determined by dividing the outer loop training data into 10 additional parts, performing an inner loop cross-validation, then selecting the shrinkage parameter which minimizes inner loop cross-validation error. Each of the 10 LASSO models in the outer loop will thus contain different shrinkage parameters, different coefficients, and potentially a different set of predictors. We will end up with a set of predicted ”pre-validated” COVID-19 probabilities for all individuals. Since the probabilities are generated from different models, the area under the curve (AUC) will be an evaluation of the model-generating process rather than of a specific model.

### RESULTS

#### The time-dependent model for individual probability of symptomatic COVID-19 in COVID Symptom Study Sweden

The AUC for the time-dependent model was 0.84 (95% CI 0.83–0.85) for the training period (April 29 and December 31, 2020), and 0.72 (95% CI 0.69–0.75) during the evaluation period (January 1 to February 10, 2021). The time-dependent model was also validated in CRUSH Covid with an AUC of 0.75 (95% CI 0.70–0.79).

**Supplementary Figure 1.** Participation rate in COVID Symptom Study Sweden per 100,000 inhabitants (≥18 years). N total =143,531.

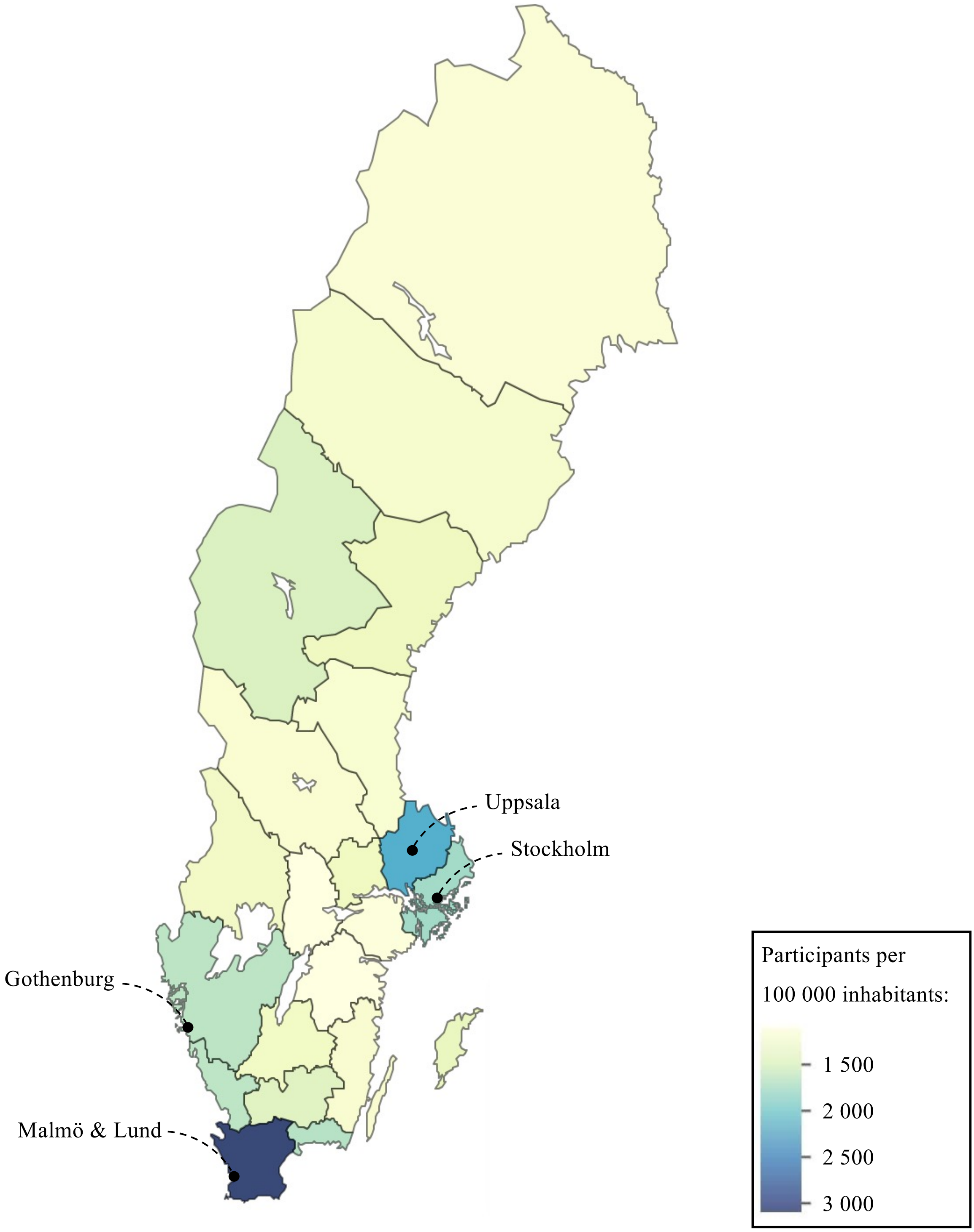

**Supplementary Figure 2.** Comparison of prevalence of different symptoms measured in NOVUS (blue) and CSSS (red) over time, with 95% confidence interval. NOVUS symptom questions were posed to representative NOVUS Sweden panelists and refer to symptoms experience the previous 14 days, while CSSS questions were posed only to those who did not feel as healthy as they normally did and refer to a single reporting day. Panel a) shows non-adjusted prevalence and panel b) prevalence of symptoms when July 1, 2020 is set to 0 for each symptom.

**a)**

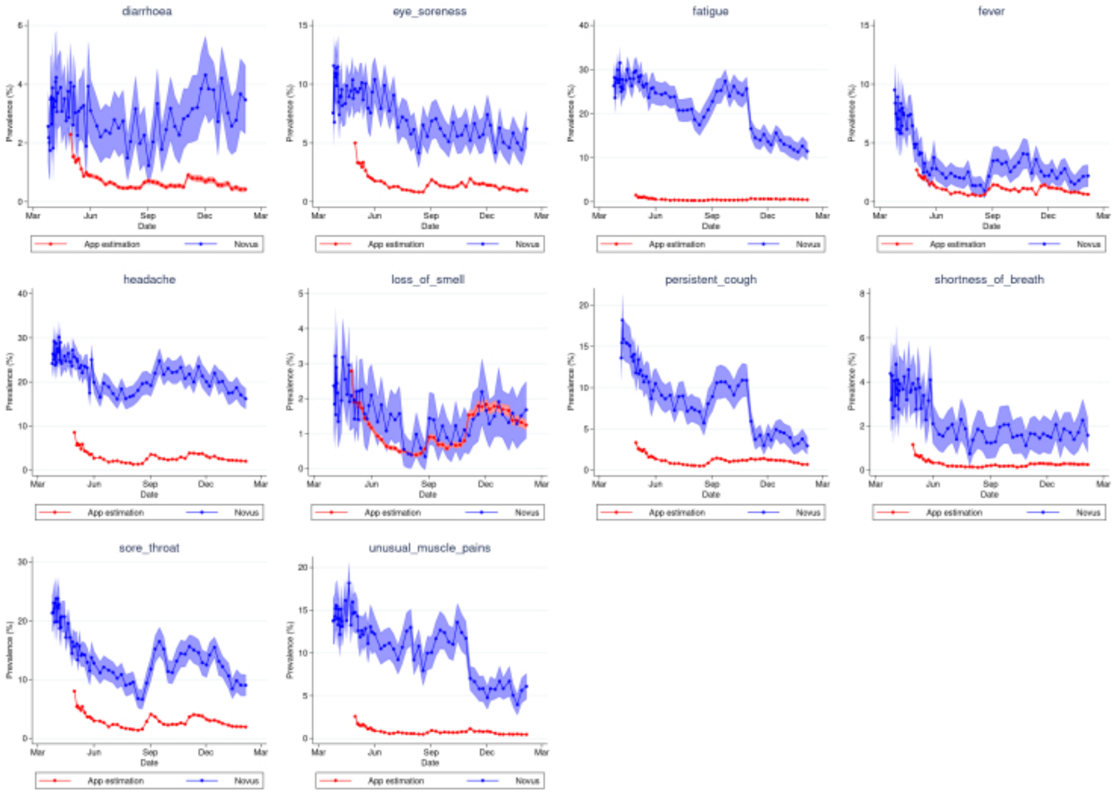

b)

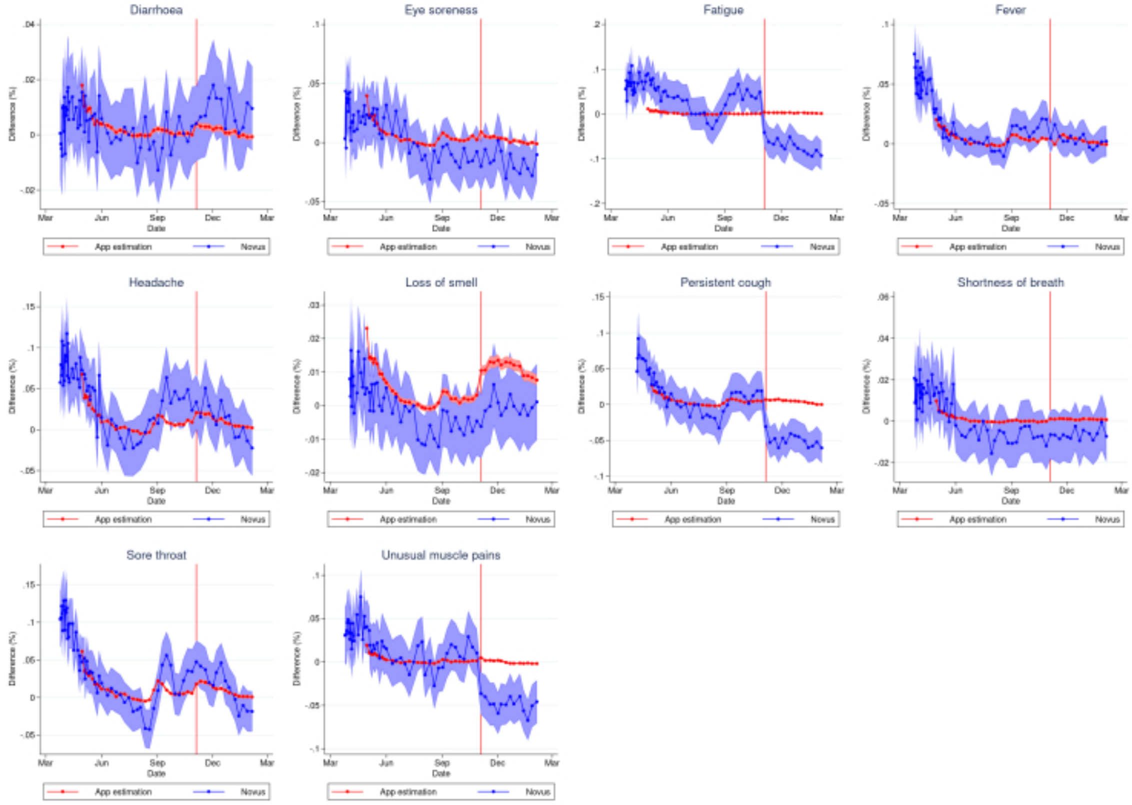
Diarrhoea
Eye soreness
Fatigue
Fever
Headache
Loss of smell
Persistent cough
Shortness of breath
Sore throat
Unusual muscle pains
Difference (%)
Date
Mar
Jun
Sep
Dec
App estimation
Novus

**Supplementary Figure 3.** Calibration plots of COVID Symptom Study Sweden main model predicting symptomatic COVID-19 (left panels) and time-dependent model (right panels); a and b) during the time period used for model training (April 29 to December 31, 2020), c and d) during the time period not used for model training (January 1 to February 10, 2021) and e and f) in CRUSH Covid data.

a)

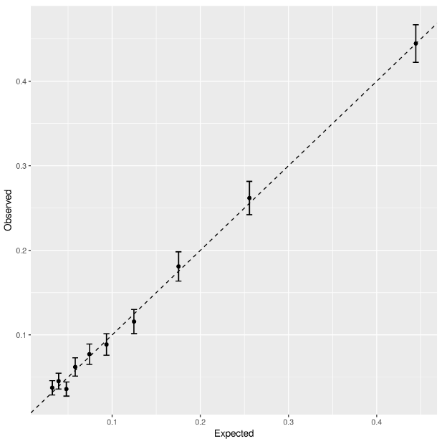

b)

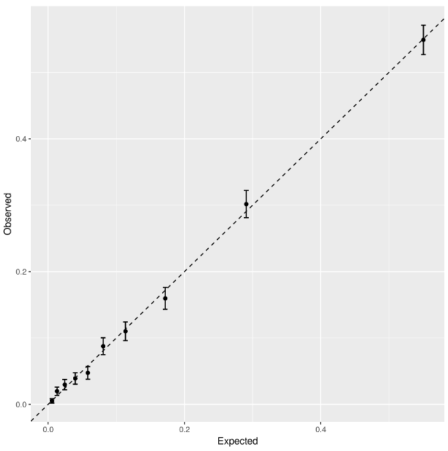

c)

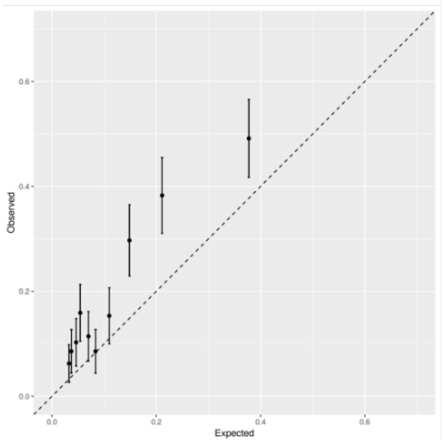

d)

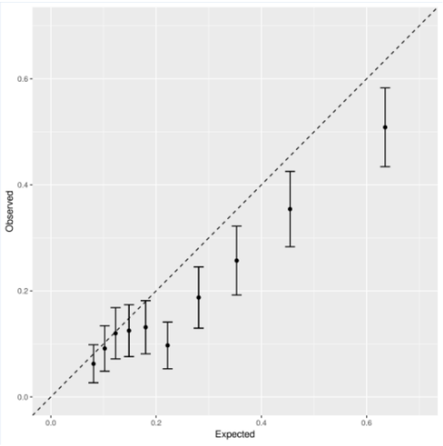

e)

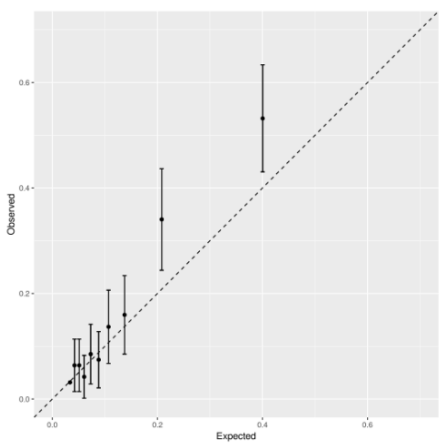

f)

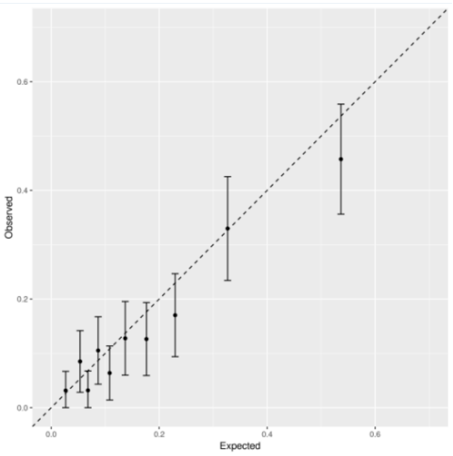

**Supplementary Figure 4.** Regional prevalence estimates, with 95% confidence interval, of symptomatic COVID-19 in COVID Symptom Study Sweden (main model, red line) in the five most populated counties in Sweden (ordered by total population ≥18 years), combined with retrospective data on a) daily number of new hospital admissions registered in the National Patient Register per 100,000 inhabitants ≥18 years, and b) daily number of new COVID-19 cases registered in SmiNet per 100,000 inhabitants ≥18 years.

**a)**

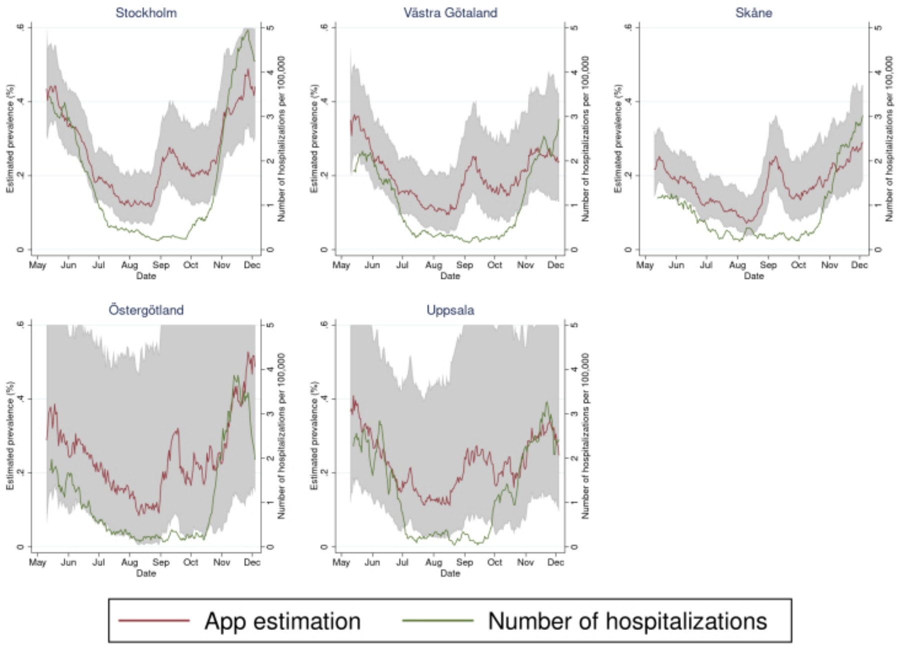

**b)**

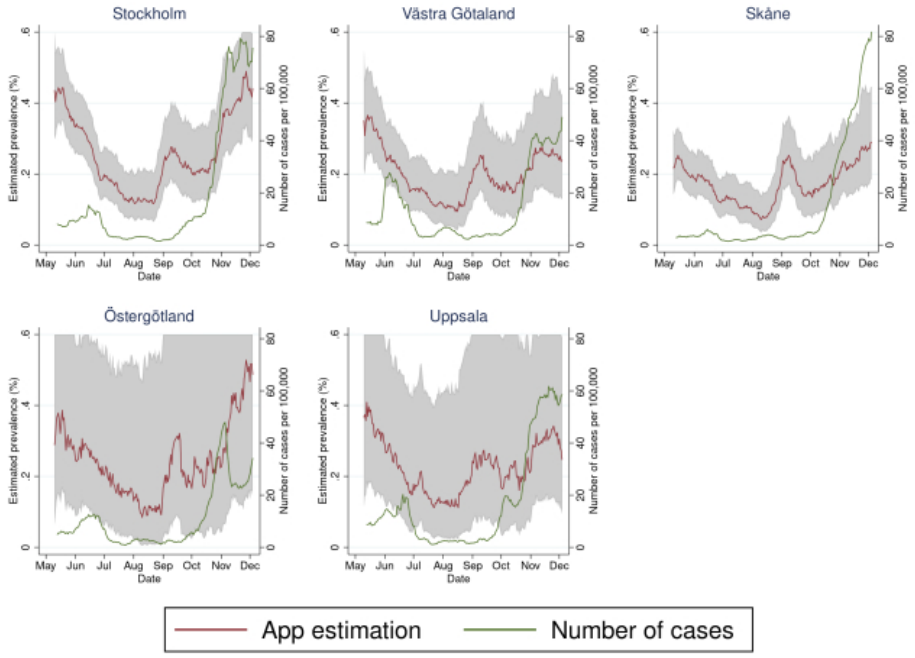

**Supplementary Figure 5. National prevalence estimates of symptomatic COVID-19 in COVID Symptom Study Sweden, depicting the main model (utilized for real-time prediction estimates) and the retrospective time-dependent model, stratified by a) sex and age (18–39, 40–64, and ≥65 years), and b) sex and age (18–39, 40–64, and ≥65 years) and healthcare professional (HP). Panel a) is further combined with retrospective data on daily number of new COVID-19 cases registered in SmiNet, per 100,000 inhabitants ≥18 years**

a)

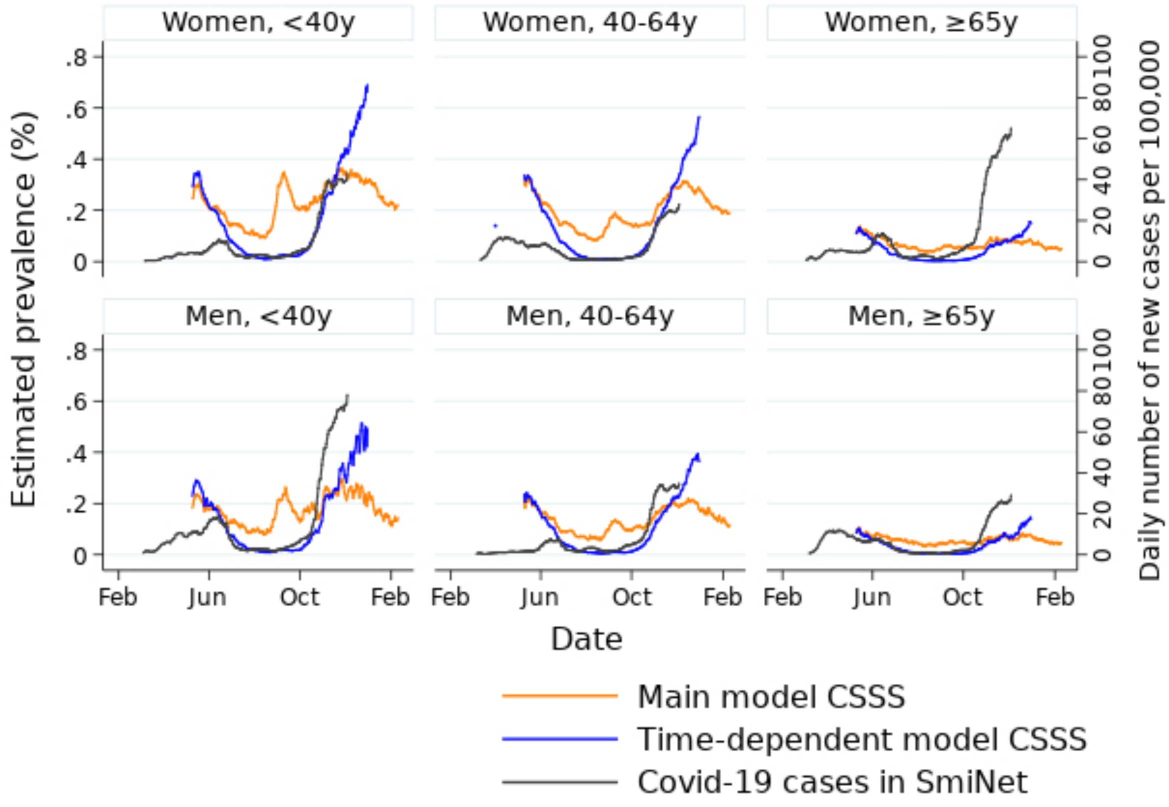

**b)**

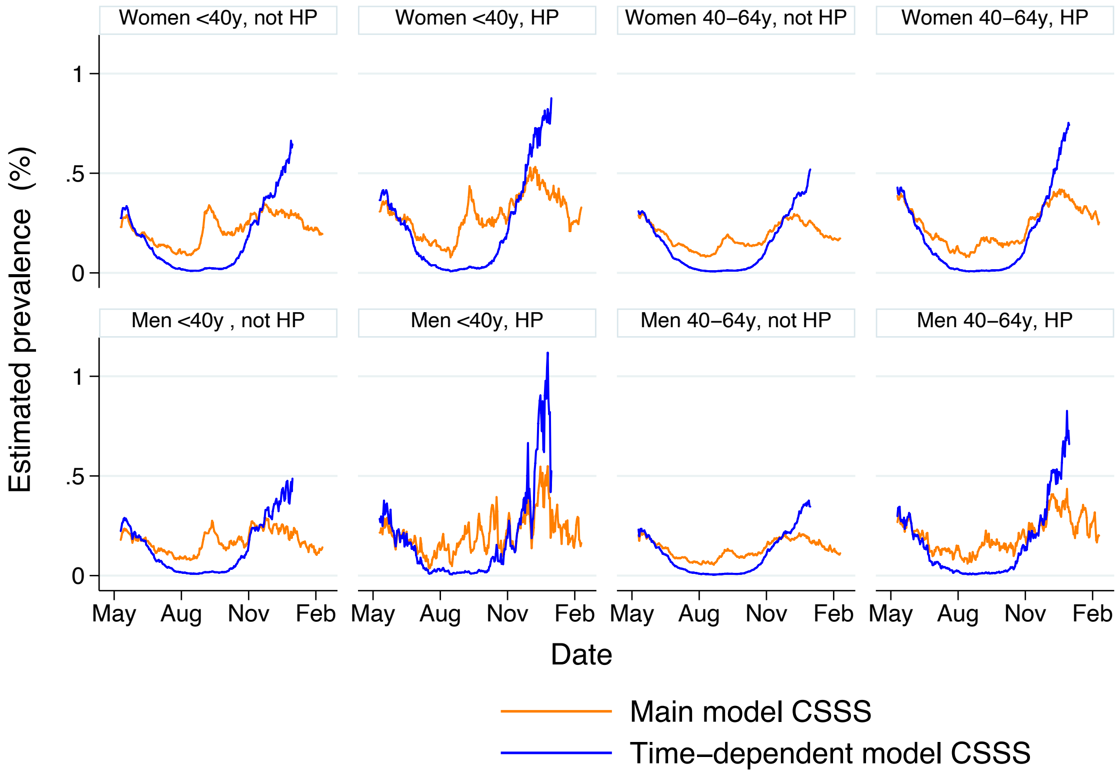

Women <40y, not HP
Women <40y, HP
Women 40–64y, not HP
Women 40–64y, HP
Men <40y , not HP
Men <40y, HP
Men 40–64y, not HP
Men 40–64y, HP
Estimated prevalence (%)
1
.5
0
May
Aug
Nov
Feb
Date
Main model CSSS
Time–dependent model CSSS

**Supplementary Figure 6. Predicted number of daily hospital admissions seven days ahead across the 16 less populated regions in Sweden (alphabetical order). The median absolute percentage errors (MdAPEs) of the predictions are denoted for the first pandemic wave (May 11 to July 3, 2020), the summer period (July 4 to October 18, 2020), and the second pandemic wave (October 19 to November 29, 2020).**

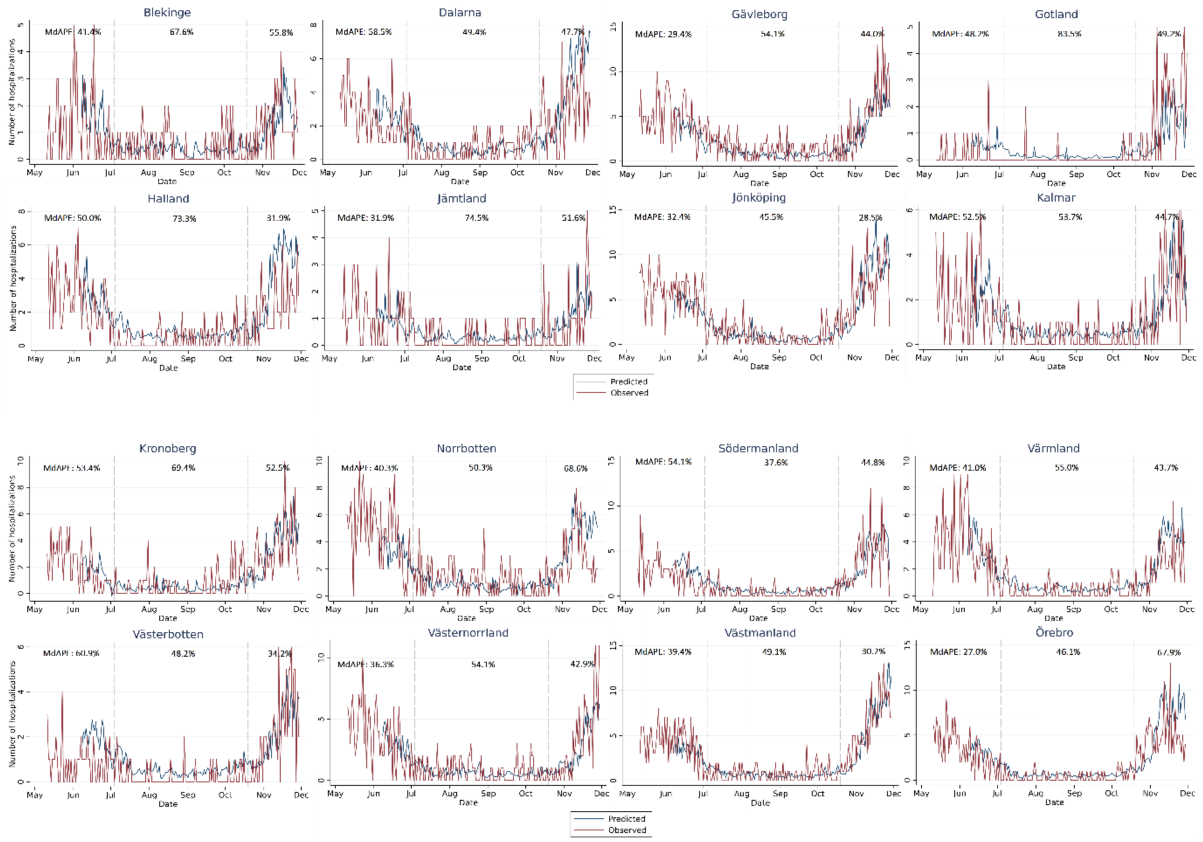

**Supplementary Figure 7. Relative error in % per day and region in hospital admission predictions over the range of observed admissions, in a) Sweden (May 11 to November 29, 2020), and b) England (April 27, 2020 to February 7, 2021).**

**a)**

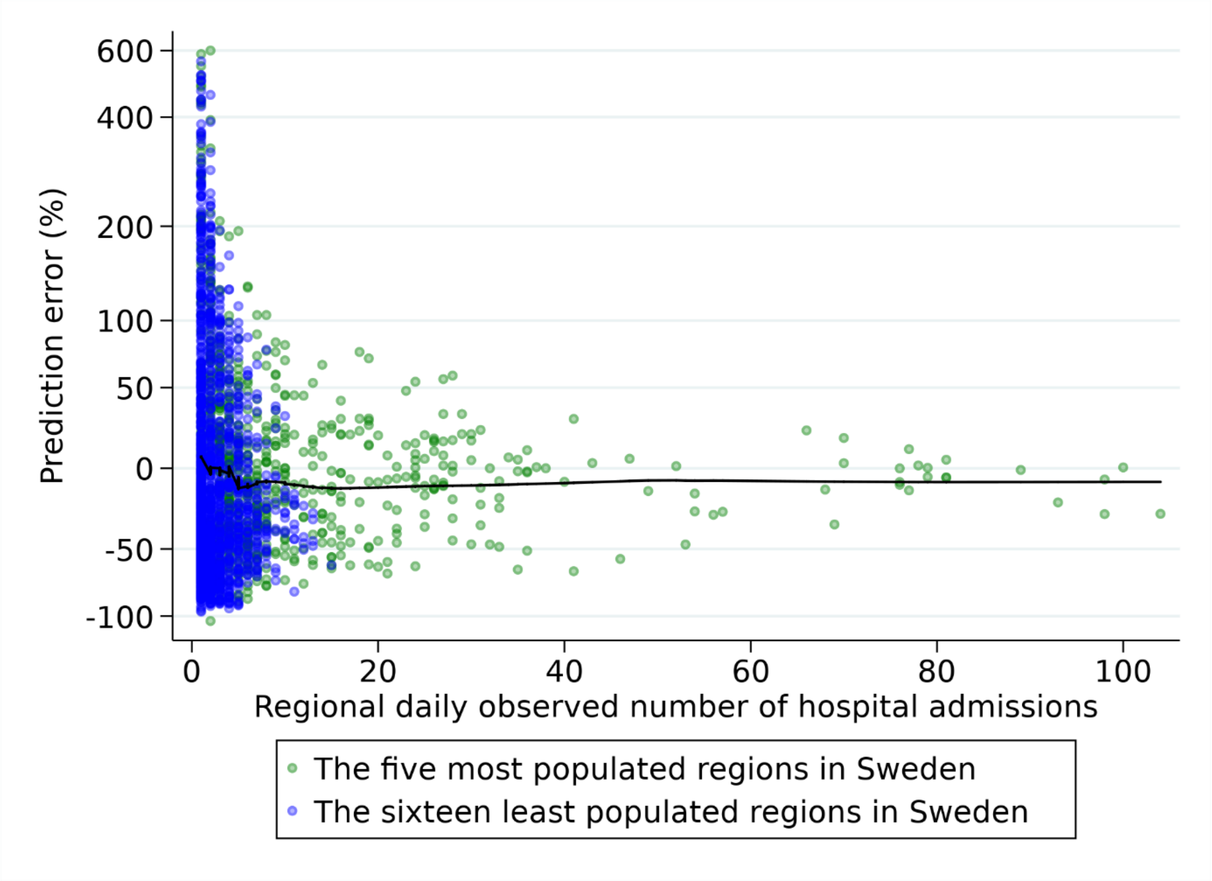

**b)**

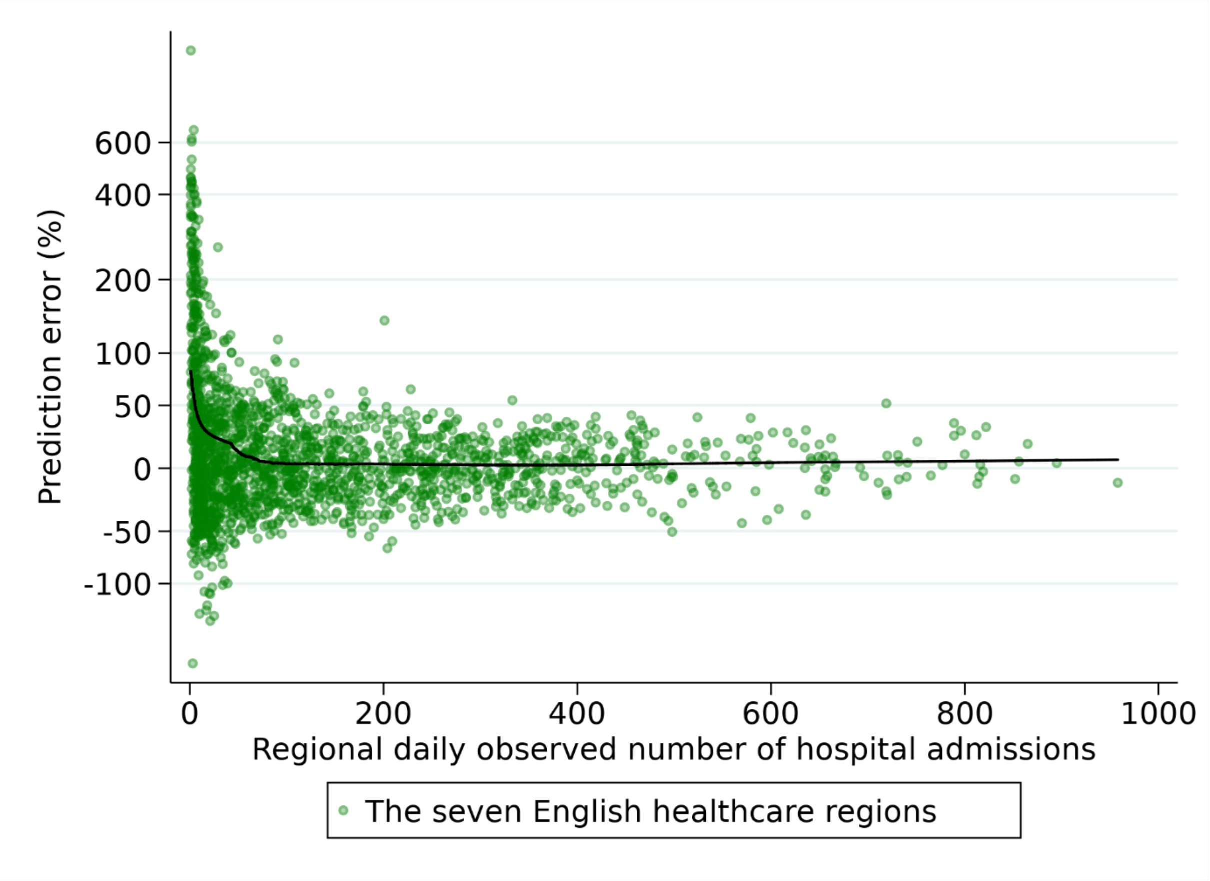
Prediction error (%)
600
400
200
100
50
0
-50
-100
0
200
400
600
800
1000
Regional daily observed number of hospital admissions
The seven English healthcare regions

**Supplementary Figure 8. Proportion of CSSS study participants reporting any symptom within seven days of joining and thereafter, respectively.**

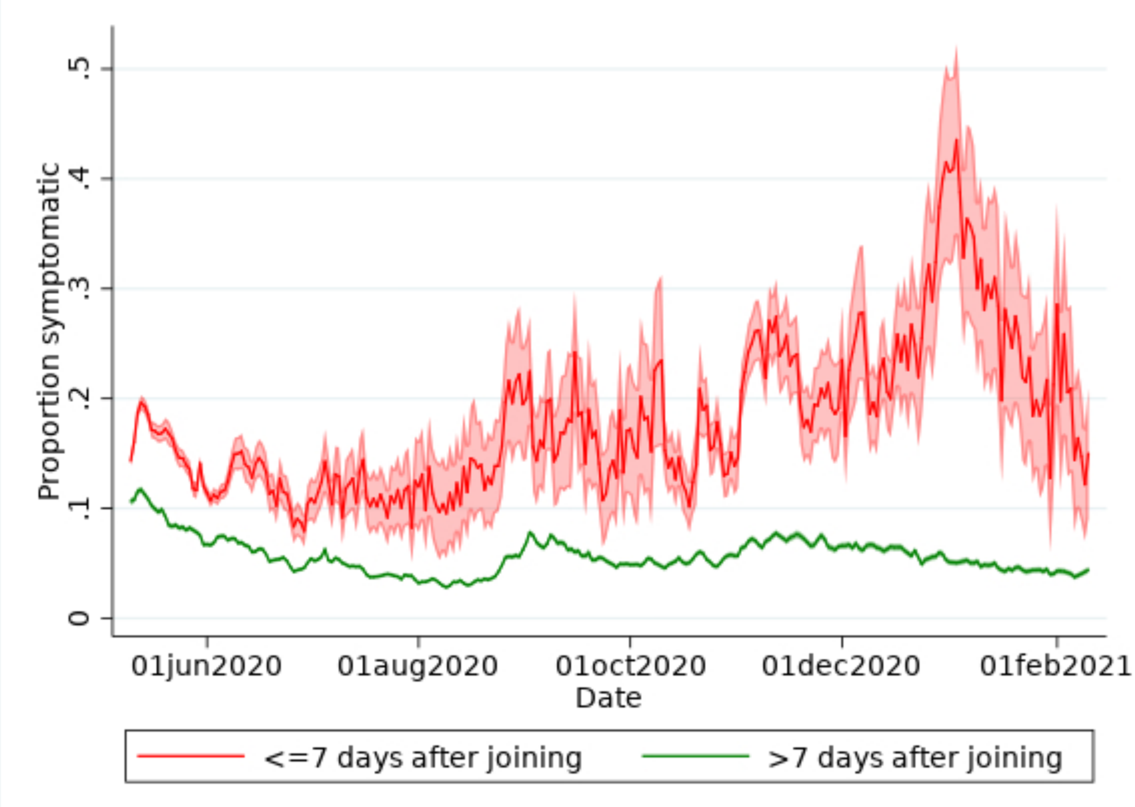

**Supplementary Table 1. COVID Symptom Study Sweden participation rate across the 21 Swedish regions.**

| Region | **Inhabitants ≥18 years** | **CSSS participants** | **CSSS participation rate per 100,000 inhabitants ≥18 years** |
|---|---|---|---|
| Blekinge | 127,494 | 2,200 | 1,726 |
| Dalarna | 229,129 | 2,793 | 1,219 |
| Gävleborg | 230,045 | 2,822 | 1,227 |
| Gotland | 48,529 | 679 | 1,399 |
| Halland | 258,093 | 4,368 | 1,692 |
| Jämtland | 104,921 | 1,603 | 1,528 |
| Jönköping | 284,392 | 3,769 | 1,325 |
| Kalmar | 193,404 | 2,395 | 1,238 |
| Kronoberg | 157,512 | 2,332 | 1,481 |
| Norrbotten | 203,368 | 2,405 | 1,183 |
| Skåne | 1,078,461 | 33,253 | 3,083 |
| Södermanland | 233,825 | 2,691 | 1,151 |
| Stockholm | 1,850,703 | 33,459 | 1,808 |
| Uppsala | 301,486 | 6,841 | 2,269 |
| Värmland | 228,081 | 2,969 | 1,302 |
| Västerbotten | 217,230 | 2,707 | 1,246 |
| Västernorrland | 195,163 | 2,621 | 1,343 |
| Västmanland | 217,652 | 2,896 | 1,331 |
| Västra Götaland | 1,363,934 | 23,186 | 1,700 |
| Örebro | 240,211 | 2,659 | 1,107 |
| Östergötland | 369,506 | 4,100 | 1,110 |

**Supplementary Table 2. SmiNet-based predicted number of daily hospital admissions seven days ahead across the 21 Swedish regions. The median absolute percentage errors (MdAPEs) of the predictions are denoted for the first pandemic wave (May 11 to July 3, 2020), the summer period (July 4 to October 18, 2020), and the second pandemic wave (October 19 to November 29, 2020).**

| | **Median absolute percentage errors (%)** | | |
|---|---|---|---|
| | First wave<br>May 11-July 3, 2020 | Summer period<br>July 4 to October 18, 2020 | Second wave<br>October 19 to November 29, 2020 |
| **All 21 regions combined** | 38.7 | 49.2 | 38.5 |
| **Top 5 most populated regions*** | 30.3 | 38.5 | 25.9 |
| **Blekinge** | 31.7 | 69.2 | 47.5 |
| **Dalarna** | 46.5 | 51.5 | 46.9 |
| **Gotland** | 71.9 | 85.8 | 45.5 |
| **Gävelborg** | 27.4 | 55.9 | 38.3 |
| **Halland** | 42.4 | 40.3 | 59.4 |
| **Jämtland** | 32.5 | 74.7 | 54.4 |
| **Jönköping** | 32.1 | 47.5 | 33.6 |
| **Kalmar** | 59.3 | 61.2 | 47.8 |
| **Kronoberg** | 53.0 | 62.6 | 48.5 |
| **Norrbotten** | 40.6 | 53.5 | 56.3 |
| **Skåne** | 51.0 | 43.2 | 23.0 |
| **Stockholm** | 13.5 | 28.7 | 24.0 |
| **Södermanland** | 44.9 | 39.0 | 45.8 |
| **Uppsala** | 36.0 | 50.9 | 25.9 |
| **Värmland** | 56.2 | 53.5 | 39.6 |
| **Västerbotten** | 45.6 | 33.6 | 34.4 |
| **Västernorrland** | 52.4 | 36.4 | 38.7 |
| **Västmanland** | 49.6 | 29.1 | 34.2 |
| **Västra Götaland** | 34.2 | 41.4 | 25.0 |
| **Örebro** | 31.2 | 51.6 | 55.1 |
| **Östergötland** | 70.7 | 38.8 | 39.0 |

* Stockholm, Västra Götaland, Skåne, Östergötland, Uppsala

**Supplementary Table 3. Study population characteristics in COVID Symptom Study England.**

| | | All | Women | Men |
|---|---|---|---|---|
| N (%)[1] | | 1,888,416 (100) | 1,239,199 (65.6) | 913,823 (34.4) |
| Age, years[2] | | 47 (35, 59) | 46 (35, 57) | 50 (38, 61) |
| | ≥65 (%) | 277,118 (14.7) | 155,573 (12.6) | 121,545 (18.7) |
| Pregnant (%) | | - | 13,394 (1.1) | - |
| BMI, kg/$m^2$ [2] | | 26 (23, 29) | 25 (22, 29) | 26 (24, 29) |
| | Obese, BMI ≥30 kg/$m^2$ (%) | 415,179 (22.0) | 281,647 (22.7) | 133,532 (20.6) |
| Current smoker (%) | | 84,036 (8.5) | 53,386 (8.5) | 30,650 (8.4) |
| Cardiovascular disease (%) | | 45,920 (2.4) | 18,663 (1.5) | 27,257 (4.2) |
| Antihypertensive medication (%) | | 160,698 (8.7) | 81,885 (6.8) | 78,813 (12.4) |
| Kidney disease (%) | | 12,961 (0.7) | 8,027 (0.6) | 4,934 (0.8) |
| Diabetes mellitus (%) | | | | |
| | Yes, type 1 | 8,861 (0.5) | 5,032 (0.4) | 3,829 (0.6) |
| | Yes, type 2 | 40,304 (2.1) | 18,140 (1.5) | 22,164 (3.4) |
| | Yes, gestational | 171 (<1) | 167 (<1) | 4 (<1) |
| | Yes, other | 1,136 (0.1) | 683 (0.1) | 453 (0.1) |
| | Yes, type not specified | 10,696 (0.6) | 5,103 (0.4) | 5,593 (0.9) |
| Lung disease (%) | | | | |
| | Yes, asthma only | 190,817 (10.1) | 133,602 (10.8) | 57,215 (8.8) |
| | Yes, both asthma and lung disease | 9,878 (0.5) | 6,729 (0.5) | 3,149 (0.5) |
| | Yes, lung disease only | 14,368 (0.8) | 8,475 (0.7) | 5,893 (0.9) |
| | Yes, type not specified | 42,006 (2.2) | 28,929 (2.3) | 13,077 (2.0) |
| Current cancer (%) | | 13,865 (1.4) | 6,722 (1.1) | 7,143 (2.0) |
| Immunosuppressive medication[3] (%) | | 63,669 (3.4) | 43,298 (3.5) | 20,371 (3.1) |
| Healthcare professional (%) | | | | |
| | Interacts with patients | 37,966 (4.1) | 31,579 (5.5) | 6,387 (1.8) |
| | Does not interact with patients | 25,702 (2.8) | 20,509 (3.5) | 5,193 (1.5) |
| Months entering the study (%) | | | | |
| | March 2020 | 1,014,687 (53.7) | 692,095 (55.9) | 322,592 (49.7) |
| | April-May 2020 | 568,030 (30.1) | 349,889 (28.2) | 218,141 (33.6) |
| | June-July 2020 | 110,298 (5.8) | 73,377 (5.9) | 36,921 (5.7) |
| | August-September 2020 | 86,506 (4.6) | 57,134 (4.6) | 29,372 (4.5) |
| | October-November 2020 | 61,011 (3.2) | 37,922 (3.1) | 23,089 (3.6) |
| | December 2020-January 2021 | 41,540 (2.2) | 24,981 (2.0) | 16,559 (2.6) |
| | February 2021 | 6,344 (0.3) | 3,801 (0.3) | 2,543 (0.4) |
| Duration of study participation, days[4]* | | 182 (62, 305) | 184 (64, 305) | 178 (60, 305) |

[1]Row percentage, [2]Median (first and third quartile), [3]Corticosteroids, methotrexate and/or biological agents (treatment of cancer and/or rheumatic disease), [4]From first to last daily report. BMI: Body Mass Index

**Supplementary Table 4. COVID Symptom Study England participation rate across the seven English healthcare regions.**

| **English healthcare region** | **Inhabitants ≥18 years** | **CSSS participants** | CSSS participation rate per 100,000 inhabitants ≥18 years |
|---|---|---|---|
| East of England | 4,787,130 | 232,430 | 4,855 |
| London | 6,775,147 | 328,087 | 4,843 |
| Midlands | 8,259,089 | 267,110 | 3,234 |
| North East and Yorkshire | 6,308,756 | 204,245 | 3,237 |
| North West | 5,632,066 | 181,538 | 3,223 |
| South East | 7,034,905 | 437,217 | 6,215 |
| South West | 4,422,690 | 238,919 | 5,402 |

**Supplementary Table 5. COVID Symptom Study England (CSSS England) median absolute percentage errors (MdAPEs) for prediction of new daily hospitalizations, across the first pandemic wave (April 6 to June 19, 2020), the summer period (June 20 to September 19, 2020), and the second pandemic wave (September 20, 2020 to February 7, 2021). The prediction model used current regional CSS England prevalence estimates and hospital data to predict hospital admissions seven days ahead.**

| | **Median absolute percentage errors (%)** | | |
|---|---|---|---|
| | First wave<br>April 6 to June 19, 2020 | Summer period<br>June 20 to September 19, 2020 | Second Wave<br>September 20, 2020 to February 7, 2021 |
| **All seven regions combined** | 22.3 | 36.0 | 19.0 |
| **East of England** | 24.6 | 40.6 | 20.8 |
| **London** | 38.8 | 25.9 | 20.5 |
| **Midlands** | 16.1 | 32.5 | 14.0 |
| **North East and Yorkshire** | 13.8 | 26.0 | 14.4 |
| **North West** | 18.7 | 35.8 | 17.5 |
| **South East** | 21.4 | 33.7 | 24.5 |
| **South West** | 33.0 | 79.4 | 22.1 |

**Supplementary Table 6. All variables in COVID Symptom Study Sweden including information on edits, additions and removals of questions during the study period April 29, 2020 to February 10, 2021, a) Baseline information, b) COVID-19 tests, and c) Daily reports.**

a) **Baseline information**

| App Display Text Translated from Swedish | | Available at launch April 29, 2020 | Date Added | Date Removed |
|---|---|---|---|---|
| **About Your Work** | | | | |
| Are you a healthcare worker (including hospital, elderly care or in the community)? | | Y | .. | .. |
| | Yes, I currently interact with patients | Y | .. | .. |
| | Yes, but I do not currently interact with patients | Y | .. | .. |
| | No | | .. | .. |
| Do you care for multiple people in the community, with direct contact with your patients? | | Y | .. | .. |
| Since the COVID-19 epidemic began, have you physically worked in? | | Y | .. | .. |
| | Hospital inpatient | Y | .. | .. |
| | Hospital outpatient | Y | .. | .. |
| | Clinic outside a hospital | Y | .. | .. |
| | Nursing home/elderly care or group care facility | Y | .. | .. |
| | Home health | Y | .. | .. |
| | School clinic | Y | .. | .. |
| | Other healthcare facility | Y | .. | .. |
| Have you EVER interacted (in person) with patients with documented or presumed COVID-19 infection? | | Y | .. | .. |
| | Yes, documented COVID-19 cases only | Y | .. | .. |
| | Yes, suspected COVID-19 case only | Y | .. | .. |
| | Yes, both documented & suspected COVID-19 cases | Y | .. | .. |
| | Not that I know of | Y | .. | .. |

| | | | | |
|---|---|---|---|---|
| Since the COVID-19 epidemic began, have you used personal protective equipment (PPE) at work?<br><br>*Depending on your specific work requirements, PPE might include gloves, masks, face shields, etc | | Y | .. | .. |
| | Always | Y | .. | .. |
| | Sometimes | Y | .. | .. |
| | Never | Y | .. | .. |
| Is the PPE you need available? | | Y | .. | .. |
| | If always: | | .. | .. |
| | I have had all the PPE I need for work | Y | .. | .. |
| | I had to reuse PPE because of shortage | Y | .. | .. |
| | If sometimes: | | .. | .. |
| | I haven't always needed to use PPE, but have had enough when I did | Y | .. | .. |
| | I would have used PPE all the time, but I haven't had enough | Y | .. | .. |
| | I've had to reuse PPE because of shortage | Y | .. | .. |
| | If never: | | .. | .. |
| | I haven't needed PPE | Y | .. | .. |
| | I needed PPE, but it wasn't available | Y | .. | .. |
| About You | | | | |
| What year were you born? | | Y | .. | .. |
| What sex were you assigned at birth? | | Y | .. | .. |
| | Female | Y | .. | .. |
| | Male | Y | .. | .. |
| | Prefer not to say | Y | .. | .. |
| | Intersex | Y | .. | .. |
| What gender do you most identify with? | | Y | .. | .. |
| | Male | Y | .. | .. |
| | Female | Y | .. | .. |

| | | | | |
|---|---|---|---|---|
| | Transgender | .. | .. | .. |
| | Do not identify as female, male or transgender | .. | .. | .. |
| | Prefer not to say | Y | .. | .. |
| Your height? | | Y | .. | .. |
| Your weight? | | Y | .. | .. |
| Your postcode? | | Y | .. | .. |
| Are you temporarily reporting from a different location than above? | | N | 2020-08-25 | .. |
| Please select which country you are now in: | | N | 2020-08-25 | .. |
| Have you EVER been exposed to someone with documented or presumed COVID-19 infection (such as co-workers, family members, or others)? Please check all that apply. | | Y | .. | .. |
| | Yes, documented COVID-19 cases | Y | .. | .. |
| | Yes, both documented & suspected COVID-19 cases | Y | .. | .. |
| | Yes, presumed COVID-19 cases | Y | .. | .. |
| | Not that I know of | Y | .. | .. |
| In general, do you have any health problems that require you to stay at home? | | Y | .. | .. |
| Do you need someone to help you on a regular basis? | | Y | .. | .. |
| If you need help, can you count on someone close to you? | | Y | .. | .. |
| Do you regularly use a stick, walking frame or wheelchair to get about? | | Y | .. | .. |
| About Your Health | | | | |
| In general, do you have any health problems that require you to limit your activities? | | Y | .. | .. |
| Are you pregnant? | | Y | 2020-03-29 | .. |
| How many weeks pregnant are you? | | N | 2020-05-07 | .. |
| At what age did your periods stop? | | N | 2020-05-07 | 2020-11-06 |
| Do your periods usually occur? | | N | 2020-05-07 | 2020-11-06 |

| | | | | |
|---|---|---|---|---|
| | Regularly every 3-6 weeks | N | 2020-05-07 | 2020-11-06 |
| | Regularly, but less often than every 6 weeks | N | 2020-05-07 | 2020-11-06 |
| | At irregular intervals | N | 2020-05-07 | 2020-11-06 |
| Are you currently having periods? | | N | 2020-05-07 | 2020-11-06 |
| | I've never had periods | N | 2020-05-07 | 2020-11-06 |
| | 'm currently having periods | N | 2020-05-07 | 2020-11-06 |
| | I've stopped having periods | N | 2020-05-07 | 2020-11-06 |
| | I'm pregnant | N | 2020-05-07 | 2020-11-06 |
| | Not currently | N | 2020-05-07 | 2020-11-06 |
| | Prefer not to say | N | 2020-05-07 | 2020-11-06 |
| How old were you when your periods stopped? | | N | .. | .. |
| Are you taking any of the following forms of hormone treatment? | | N | 2020-05-07 | 2020-11-06 |
| | No | N | 2020-05-07 | 2020-11-06 |
| | Combined oral contraceptive pill | N | 2020-05-07 | 2020-11-06 |
| | Progesterone only pill | N | 2020-05-07 | 2020-11-06 |
| | Mirena or other hormone coil | N | 2020-05-07 | 2020-11-06 |
| | Depot injection or implant | N | 2020-05-07 | 2020-11-06 |
| Hormone Replacement Therapy | | N | 2020-05-07 | 2020-11-06 |
| | Estrogen hormone therapy due to symptoms related to menopause | N | 2020-05-07 | 2020-11-06 |
| | Estrogen hormone therapy for gender transitioning | N | 2020-05-07 | 2020-11-06 |
| | Testosterone hormone therapy | N | 2020-05-07 | 2020-11-06 |
| | Prefer not to say | N | 2020-05-07 | 2020-11-06 |
| | Other | N | 2020-05-07 | 2020-11-06 |
| Do you have heart disease? | | Y | .. | .. |
| Do you have diabetes? | | Y | .. | .. |
| What kind of diabetes do you have? | | N | 2020-06-19 | .. |
| | Type 1 Diabetes | N | 2020-06-19 | .. |
| | Type 2 Diabetes | N | 2020-06-19 | .. |

| | | | | |
|---|---|---|---|---|
| | Gestational Diabetes (during pregnancy) | N | 2020-06-19 | .. |
| | Unsure | N | 2020-06-19 | .. |
| | Prefer not to say | N | 2020-06-19 | .. |
| | Other, please specify | N | 2020-06-19 | .. |
| What was your most recent hemoglobin A1c? | | N | 2020-06-19 | .. |
| What year were you diagnosed with diabetes? | | N | 2020-06-19 | .. |
| Which diabetes treatments do you use? Select all that apply | | N | 2020-06-19 | .. |
| | None | N | 2020-06-19 | .. |
| | Lifestyle modification (e.g. exercise, diet, weight management) | N | 2020-06-19 | .. |
| | Once or twice daily injections of long acting<br>insulin or \"basal\" insulin (e.g. glargine, detemir, degludec or NPH in premixed insulin) | N | 2020-06-19 | .. |
| | Rapid acting insulin injections with meals or as \"bolus\" (e.g. lispro, aspart, glulisine, or in premixed insulin) | N | 2020-06-19 | .. |
| | Continuous infusion in an insulin pump | N | 2020-06-19 | .. |
| | Injections of a non-insulin diabetes medication or GLP-1 receptor agonists (e.g. semaglutide, dulaglutide, liraglutide) | N | 2020-06-19 | .. |
| | Medications taken by mouth | N | 2020-06-19 | .. |
| | Prefer not to say | N | 2020-06-19 | .. |
| Which oral medications do you use? Select all that apply | | N | 2020-06-19 | .. |
| | Biguanide (Metformin) | N | 2020-06-19 | .. |
| | Sulfonylurea (e.g. glipizide, glyburide, glimepiride, gliclazide) | N | 2020-06-19 | .. |
| | DPP-4 inhibitors (e.g. saxagliptin, sitagliptin, linagliptin) | N | 2020-06-19 | .. |
| | Meglitinides (e.g. repaglinide, nateglinide) | N | 2020-06-19 | .. |
| | Thiazolidinediones or Glitazones (e.g. rosiglitazone, pioglitazone) | N | 2020-06-19 | .. |

| | | | | |
|---|---|---|---|---|
| | SGLT-2 inhibitors (e.g. canagliflozin, dapagliflozin) | N | 2020-06-19 | .. |
| | Other oral diabetes medication not listed | N | 2020-06-19 | .. |
| Do you use a continuous glucose monitor (CGM)? | | N | 2020-06-19 | .. |
| Do you have lung disease or asthma? | | Y | .. | .. |
| Do you have lung disease? | | N | 2020-06-05 | .. |
| Do you have asthma? | | N | 2020-06-05 | .. |
| Do you have hayfever? | | N | 2020-06-05 | .. |
| Do you have eczema? | | N | 2020-06-05 | .. |
| Do you smoke? | | Y | .. | .. |
| | Yes | Y | .. | .. |
| | Not currently | Y | .. | .. |
| | Never | Y | .. | .. |
| How many years since you last smoked? | | Y | .. | .. |
| Do you have kidney disease? | | Y | .. | .. |
| Are you living with cancer? | | Y | .. | .. |
| What type of cancer do you have? | | Y | .. | .. |
| Are you on chemotherapy or immunotherapy for cancer? | | Y | .. | .. |
| Do you regularly take immunosuppressant medications (including steroids, methotrexate, biologics)? | | Y | .. | .. |
| Do you regularly take aspirin (baby aspirin or standard dose)? | | Y | .. | .. |
| Do you regularly take "NSAIDs" like ibuprofen, nurofen, diclofenac, naproxen? | | Y | | |
| Are you regularly taking blood pressure medications ending in -pril, such as enalapril, lisinopril, captopril, ramipril) ? | | Y | | |
| Are you regularly taking blood pressure medications ending in -sartan, such as losartan, valsartan, irbesartan? | | Y | | |
| Are you regularly taking any blood pressure medications? | | Y | | |
| Have you been taking any vitamins or other supplements regularly for more than 3 | | N | 2020-06-02 | 2020-11-06 |

| | | | | |
|---|---|---|---|---|
| months? Regularly means more than 3 times a week on average. Select all that apply. | | | | |
| | No | N | 2020-06-02 | 2020-11-06 |
| | Vitamin C | N | 2020-06-02 | 2020-11-06 |
| | Vitamin D | N | 2020-06-02 | 2020-11-06 |
| | Omega-3 or Fish Oil | N | 2020-06-02 | 2020-11-06 |
| | Zinc | N | 2020-06-02 | 2020-11-06 |
| | Garlic | N | 2020-06-02 | 2020-11-06 |
| | Probiotics | N | 2020-06-02 | 2020-11-06 |
| | Multi-vitamins and minerals | N | 2020-06-02 | 2020-11-06 |
| | Other, please specify | N | 2020-06-02 | 2020-11-06 |
| | Prefer not to say | N | 2020-06-02 | 2020-11-06 |
| What is your blood type? | | N | 2020-07-02 | .. |
| | Type A | N | 2020-07-02 | .. |
| | Type B | N | 2020-07-02 | .. |
| | Type AB | N | 2020-07-02 | .. |
| | Type O | N | 2020-07-02 | .. |
| | I'm unsure | N | 2020-07-02 | .. |
| | Prefer not to say | N | 2020-07-02 | .. |
| Previous Exposure to COVID | | | | |
| Have you felt unwell in the month before you started reporting on this app? | | Y | .. | .. |
| Did you have any of the following symptoms (please select all that apply): | | Y | .. | .. |
| | Loss of smell/taste | Y | .. | .. |
| | Unusual shortness of breath | Y | .. | .. |
| | Unusual fatigue | Y | .. | .. |
| | Fever | Y | .. | .. |
| | Skipped meals | Y | .. | .. |
| | Persistent cough | Y | .. | .. |
| | Diarrhoea | Y | .. | .. |

| | | | | |
|---|---|---|---|---|
| | Unusual chest pain or tightness in your chest | Y | .. | .. |
| | Hoarse Voice | Y | .. | .. |
| | Abdominal Pain | Y | .. | .. |
| | Confusion, Disorientation, Drowsiness | Y | .. | .. |
| Are you still experiencing symptoms? | | Y | .. | .. |
| How have your symptoms changed over the last few days? | | Y | .. | .. |
| | Much better | Y | .. | .. |
| | A little better | Y | .. | .. |
| | The same | Y | .. | .. |
| | A little worse | Y | .. | .. |
| | Much worse | Y | .. | .. |
| Do you think you have already had COVID-19, but were not tested? | | Y | .. | 2020-11-06 |
| Do you have the classic symptoms (high fever and persistent cough) for several days? | | Y | .. | 2020-11-06 |
| How many days ago did your symptoms start? | | Y | .. | .. |
| Healthcare Worker Exposure | | | | |
| In the last day, have you interacted with any patients? | | Y | .. | .. |
| In the last day, did you treat patients in person with documented or presumed COVID-19 infection? | | Y | .. | .. |
| | Yes, documented COVID-19 cases | Y | .. | .. |
| | Yes, both documented & suspected COVID-19 cases | Y | .. | .. |
| | Yes, presumed COVID-19 cases | Y | .. | .. |
| | Not that I know of | Y | .. | .. |
| In the last day, did you use personal protective equipment (PPE) at work? *Depending on your specific work requirements, PPE might include gloves, masks, face shields, etc. | | Y | .. | .. |
| | Always | Y | .. | .. |

| | | | | |
|---|---|---|---|---|
| | Sometimes | Y | .. | .. |
| | Never | Y | .. | .. |
| | If always: | Y | .. | .. |
| | I have had all the PPE I need for work | Y | .. | .. |
| | I had to reuse PPE because of shortage | Y | .. | .. |
| | If sometimes: | Y | .. | .. |
| | I haven’t always needed to use PPE, but have had enough when I did | Y | .. | .. |
| | I would have used PPE all the time, but I haven’t had enough | Y | .. | .. |
| | I’ve had to reuse PPE because of shortage | Y | .. | .. |
| | If never: | Y | .. | .. |
| | I haven’t needed PPE | Y | .. | .. |
| | I needed PPE, but it wasn't available | Y | .. | .. |
| Levels of Isolation (legacy) | | | | |
| How much have you been isolating over the last week? | | Y | .. | 2020-05-07 |
| | I have not left the house | Y | .. | 2020-05-07 |
| | I rarely leave the house and when I do, I have little interaction with others (e.g. for exercise) | Y | .. | 2020-05-07 |
| | I rarely leave the house but had to visit somewhere with lots of people (e.g. hospital/clinic, groceries) | Y | .. | 2020-05-07 |
| | I have to leave the house often and am in contact with other people (e.g. still working outside the house or using public transport) | Y | .. | 2020-05-07 |
| Levels of Isolation (new) | | | | |
| How much have you been isolating over the last week? | | N | 2020-05-07 | 2020-09-29 |
| In the last week, how many times have you been outside, with little interaction with people outside your household (e.g. exercise)? | | N | 2020-05-07 | 2020-09-29 |

| | | | | |
|---|---|---|---|---|
| In the last week, how many times have you visited somewhere with lots of people (e.g. groceries, public transport, work)? | | N | 2020-05-07 | 2020-09-29 |
| In the last week, how many times have you visited a healthcare provider (e.g. hospital, clinic, dentist, pharmacy)? | | N | 2020-05-07 | 2020-09-29 |
| In the last week, did you wear a face mask when outside the house? | | N | 2020-06-12 | 2020-09-29 |
| | Never | N | 2020-06-12 | 2020-09-29 |
| | Sometimes | N | 2020-06-12 | 2020-09-29 |
| | Most of the time | N | 2020-06-12 | 2020-09-29 |
| | Always | N | 2020-06-12 | 2020-09-29 |
| | Not applicable | N | 2020-06-12 | 2020-09-29 |
| What kind of face mask do you use? Check all that apply | | N | 2020-06-12 | 2020-09-29 |
| | Cloth or scarf | N | 2020-06-12 | 2020-09-29 |
| | Surgical mask | N | 2020-06-12 | 2020-09-29 |
| | N95/FFP respirator | N | 2020-06-12 | 2020-09-29 |
| | Not sure/prefer not to say | N | 2020-06-12 | 2020-09-29 |
| | Other, please specify | N | 2020-06-12 | 2020-09-29 |
| Lifestyle Screen | | | | |
| Since the start of march..... | | N | 2020-06-19 | 2020-09-29 |
| How has your weight changed? | | N | 2020-06-19 | 2020-09-29 |
| | Increased | N | 2020-06-19 | 2020-09-29 |
| | Decreased | N | 2020-06-19 | 2020-09-29 |
| | Stayed the same | N | 2020-06-19 | 2020-09-29 |
| | Prefer not to say | N | 2020-06-19 | 2020-09-29 |
| If your weight has increased/decreased: | | .. | .. | .. |
| By how much (an estimate is fine)? | | N | 2020-06-19 | 2020-09-29 |
| How was your diet changed in your opinion? | | N | 2020-06-19 | 2020-09-29 |
| | It has become healthier | N | 2020-06-19 | 2020-09-29 |
| | It has become more unhealthy | N | 2020-06-19 | 2020-09-29 |
| | It has stayed the same | N | 2020-06-19 | 2020-09-29 |

| | | | | |
|---|---|---|---|---|
| | Prefer not to say | N | 2020-06-19 | 2020-09-29 |
| How has your snacking changed? | | N | 2020-06-19 | 2020-09-29 |
| | I am snacking more | N | 2020-06-19 | 2020-09-29 |
| | I am snacking less | N | 2020-06-19 | 2020-09-29 |
| | My snacking levels are the same | N | 2020-06-19 | 2020-09-29 |
| | Prefer not to say | N | 2020-06-19 | 2020-09-29 |
| How has your alcohol consumption changed? | | N | 2020-06-19 | 2020-09-29 |
| | I don't drink alcohol | N | 2020-06-19 | 2020-09-29 |
| | I am drinking more alcohol | N | 2020-06-19 | 2020-09-29 |
| | I am drinking less alcohol | N | 2020-06-19 | 2020-09-29 |
| | My alcohol consumption is the same | N | 2020-06-19 | 2020-09-29 |
| | Prefer not to say | N | 2020-06-19 | 2020-09-29 |
| Have your physical activity levels changed? | | N | 2020-06-19 | 2020-09-29 |
| | Yes, increased | N | 2020-06-19 | 2020-09-29 |
| | Yes, decreased | N | 2020-06-19 | 2020-09-29 |
| | No change, has remained the same | N | 2020-06-19 | 2020-09-29 |
| | Prefer not to say | N | 2020-06-19 | 2020-09-29 |

**b) COVID-19 tests**

| App Display Text Translated from Swedish | | Available at launch April 29, 2020 | Date Added | Date Removed | Comment |
|---|---|---|---|---|---|
| **Covid Test (v1)** | | | | | |
| Have you had a test for COVID-19? | | Y | .. | .. | New version below |
| Did you test positive for COVID-19? | | Y | .. | .. | New version below |
| When was your test? | | N | 2020-05-01 | .. | .. |
| How was this test performed? | | Y | .. | .. | .. |
| | Someone swabbed my nose | Y | .. | .. | .. |
| | Someone swabbed my throat | Y | .. | .. | .. |
| | I spat in a cup/tube | Y | .. | .. | .. |
| | I gave a blood sample | Y | .. | .. | .. |
| | Other, please specify | Y | .. | .. | .. |
| **COVID Test Screen (v2)** | | | | | |
| How was this test performed? | | N | 2020-07-02 | .. | .. |
| | Someone swabbed my nose (Deprecated) | N | 2020-07-02 | .. | .. |
| | Someone swabbed my throat (Deprecated) | N | 2020-07-02 | .. | .. |
| | A nose or throat swab | N | | .. | .. |
| | I spat in a cup/tube | N | 2020-07-02 | .. | .. |
| | I gave a blood sample (Deprecated) | N | 2020-07-02 | .. | .. |
| | I pricked my finger and gave blood | N | 2020-07-02 | .. | .. |
| | My blood was drawn via a needle | N | 2020-07-02 | .. | .. |
| | Other, please specify | N | 2020-07-02 | .. | .. |
| Where was your test performed? | | N | 2020-07-02 | .. | .. |
| | At Home | N | 2020-07-02 | .. | .. |
| | Drop In Testcenter | N | 2020-07-02 | .. | .. |

| | | | | | |
|---|---|---|---|---|---|
| | Hospital | N | 2020-07-02 | .. | .. |
| | GP | N | 2020-07-02 | .. | .. |
| | Chemist Pharmacy (UK)<br>Store or pharmacy clinic (US) | N | 2020-07-02 | .. | .. |
| | Work | N | 2020-07-02 | .. | .. |
| | Other, please specify | N | 2020-07-02 | .. | .. |
| Did you wait less than 2 hours to get the results (i.e. was it a rapid test such as a lateral flow test?) | | N | 2020-12-23 | .. | .. |
| **Vaccine Doses** | | | | | |
| Vaccine Type | | N | 2021-02-12 | .. | .. |
| | | N | 2021-02-12 | .. | .. |
| When did you have your dose? | | N | 2021-02-12 | .. | .. |
| | | N | 2021-02-12 | .. | .. |
| Vaccine Symptoms | | | | | |
| Are you experiencing any symptoms near the injection site?<br>(Check all that apply) | | N | 2021-02-12 | .. | .. |
| | Pain | N | 2021-02-12 | .. | .. |
| | Redness | N | 2021-02-12 | .. | .. |
| | Swelling | N | 2021-02-12 | .. | .. |
| | Swollen glands in the armpit | N | 2021-02-12 | .. | .. |
| | Warmth | N | 2021-02-12 | .. | .. |
| | Itch | N | 2021-02-12 | .. | .. |
| | Tenderness | N | 2021-02-12 | .. | .. |
| | Bruising | N | 2021-02-12 | .. | .. |
| | Other (free text) | N | 2021-02-12 | .. | .. |
| **Vaccine Hesitancy** | | | | | |
| Would you accept a COVID-19 vaccine if offered? | | N | 2021-02-12 | .. | .. |
| | Yes | N | 2021-02-12 | .. | .. |
| | No | N | 2021-02-12 | .. | .. |
| | Unsure | N | 2021-02-12 | .. | .. |

| | | | | | |
|---|---|---|---|---|---|
| If "No" or "Unsure": Tell us why (tick all that apply) | | N | 2021-02-12 | .. | .. |
| | Personal belief / philosophical reasons | N | 2021-02-12 | .. | .. |
| | Pregnancy / breastfeeding | N | 2021-02-12 | .. | .. |
| | Concerned about long term side effects | N | 2021-02-12 | .. | .. |
| | Do not know enough about it | N | 2021-02-12 | .. | .. |
| | Illness / medication | N | 2021-02-12 | .. | .. |
| | Do not think it will be available to me | N | 2021-02-12 | .. | .. |
| | Do not think it is necessary | N | 2021-02-12 | .. | .. |
| | Do not think it will work | N | 2021-02-12 | .. | .. |
| | Concerned about adverse reaction | N | 2021-02-12 | .. | .. |
| | Prefer not to say | N | 2021-02-12 | .. | .. |
| | Other (free text) | N | 2021-02-12 | .. | .. |

**c) Daily reports**

| App Display Text Translated from Swedish | | Available at launch April 29, 2020 | Date Added | Date Removed |
|---|---|---|---|---|
| **How You Feel** | | | | |
| How do you feel right now? | | Y | .. | .. |
| | Other | Y | .. | .. |
| | I'm not feeling quite right | Y | .. | .. |
| Are you experiencing any of the following symptoms? | | Y | .. | .. |
| **Describe Symptoms** | | | | |
| v1: Do you have a fever or feel too hot?<br>v2 Text: Fever (at least 37.8C or 100F) | | Y | .. | .. |
| If you are able to measure it, what is your temperature? | | Y | .. | |
| Do you feel chills or shivers (feel too cold)? | | Y | .. | 2020-11-03 |
| v1: Do you have a persistent cough (coughing a lot for more than an hour, or 3 or more coughing episodes in 24 hours)?<br>v2: Persistent cough (coughing a lot for more than an hour or 3 or more coughing episodes in 24 hours) | | Y | .. | .. |
| v1: Are you experiencing unusual fatigue?<br>v2: Unusual fatigue... | | Y | .. | .. |
| | How severe is your fatigue? | Y | .. | .. |
| | Mild fatigue | Y | .. | .. |
| | Severe fatigue - I struggle to get out of bed | Y | .. | .. |
| v1: Are you experiencing unusual shortness of breath?<br>v2: Shortness of breath or trouble breathing | | Y | .. | .. |
| | How serious are your breathing difficulties? | Y | .. | .. |
| | Yes. Mild symptoms – slight shortness of breath during ordinary activity | Y | .. | .. |
| | Yes. Significant symptoms - breathing is comfortable only at rest | Y | .. | .. |
| | Yes. Severe symptoms – breathing is difficult even at rest | Y | .. | .. |
| v1: Do you have a loss of smell/taste?<br>v2: Loss of smell / taste | | Y | .. | .. |

| | | | |
|---|---|---|---|
| v1: Do you have an unusually hoarse voice?<br>v2: Unusually hoarse voice | Y | .. | .. |
| v1: Are you feeling an unusual chest pain or tightness in your chest?<br>v2: Unusual chest pain or tightness in your chest | Y | .. | .. |
| v1: Do you have an unusual abdominal pain?<br>v2: Unusual abdominal pain or stomach ache | Y | .. | .. |
| v1: Are you experiencing diarrhoea?<br>v2: Diarrhoea | Y | .. | .. |
| How many loose stools in the last 24 hours? | N | 2020-05-07 | 2020-11-03 |
| How often are you experiencing headaches? | N | 2020-05-07 | .. |
| v1: Do you have any of the following symptoms: confusion, disorientation or drowsiness?<br>v2: Confusion, disorientation or drowsiness | Y | .. | .. |
| Do your eyes have any unusual eye-soreness or discomfort (e.g. light sensitivity, excessive tears, or pink/red eye)? | Y | .. | .. |
| v1: Have you been skipping meals?<br>v2: Skipping meals | Y | .. | .. |
| v1: Do you have a headache?<br>v2: Headache | Y | .. | .. |
| How much of the day are you experiencing headaches? | Y | .. | .. |
| The entire day | Y | .. | .. |
| Most part of the day | Y | .. | .. |
| Part of the day | Y | .. | .. |
| v1: Have you felt nauseous or experienced vomiting?<br>v2: Nausea or vomiting | Y | .. | .. |
| v1: Are you experiencing dizziness or light-headedness?<br>v2: Dizziness or light-headedness | Y | .. | .. |
| v1: Do you have a sore throat?<br>v2: Sore or painful throat | Y | .. | .. |
| v1: Do you have unusual strong muscle pains?<br>v2: Unusual strong muscle pains or aches | Y | .. | .. |
| v1: Have you had raised, red, itchy welts on the skin or sudden swelling of the face or lips?<br>v2: Raised, red, itchy welts on the skin or sudden swelling of the face or lips | Y | .. | .. |

| | | | |
|---|---|---|---|
| v1: Have you had any red/purple sores or blisters on your feet, including your toes?<br>v2: Red/purple sores or blisters on your feet, including your toes | Y | .. | .. |
| Are there other important symptoms you want to share with us? | Y | .. | .. |
| v1: Are your current symptoms unusual compared to your usual hay fever or seasonal allergies?<br>v2: Increase in your usual allergy symptoms | Y | .. | .. |
| v2: Rash on your arms or torso | N | 2020-11-03 | .. |
| v2: Strange, unpleasant sensations in your skin like pins & needles or burning | N | 2020-11-03 | .. |
| v2: Unusual hair loss | N | 2020-11-03 | .. |
| v2: Feeling down, depressed or hopeless | N | 2020-11-03 | .. |
| v2: Loss of concentration or memory (brain fog) | N | 2020-11-03 | .. |
| v2: Altered smell / taste (things smell or taste different to usual) | N | 2020-11-03 | .. |
| v2: Runny nose | N | 2020-11-03 | .. |
| v2: Sneezing more than usual | N | 2020-11-03 | .. |
| v2: Earache | N | 2020-11-03 | .. |
| v2: Ringing in your ears | N | 2020-11-03 | .. |
| v2: Swollen neck glands | N | 2020-11-03 | .. |
| v2: Unusually fast or irregular heartbeat (palpitations) | N | 2020-11-03 | .. |
| Mouth or tongue ulcers | N | 2021-02-04 | .. |
| Changes to tongue surface | N | 2021-02-04 | .. |
| v2: Unusual joint pains or aches | N | 2020-12-09 | .. |
| **Where Are You** | | | |
| Where are you right now | Y | .. | .. |
| I'm at home. I haven't been to the hospital for suspected COVID-19 symptoms. | Y | .. | .. |
| I am at the hospital with suspected COVID-19 symptoms. | Y | .. | .. |
| I am back from the hospital, I'd like to tell you about my treatment. | Y | .. | .. |

| | | | | |
|---|---|---|---|---|
| | I am back from the hospital, I've already told you about my treatment. | Y | .. | .. |
| **Treatment Selection** | | | | |
| What treatment did you receive while in the hospital / What treatment are you receiving right now? | | Y | .. | .. |
| | None | Y | .. | .. |
| | Oxygen and fluids Breathing support administered through an oxygen mask, no pressure applied | Y | .. | .. |
| | Non-invasive ventilation breathing support administered through an oxygen mask, which pushes oxygen into your lungs | Y | .. | .. |
| | Invasive ventilation breathing support administered through an inserted tube. People are usually asleep for this procedure | Y | .. | .. |
| | Other | Y | .. | .. |

**Supplementary Table 7. Characteristics for participants in COVID Symptom Study Sweden reporting sex as 'intersex' or 'other'.**

| | | Intersex | Other |
|---|---|---|---|
| N (%)[1] | | 36 (15,3) | 200 (84,7) |
| Age, years[2] | | 46 (34, 55) | 38 (27, 50) |
| | ≥65 (%) | 6 (16.7) | 10 (5.0) |
| Pregnant (%) | | 1 (2.8) | 0 (0.0) |
| BMI, $kg/m^2$ [2] | | 24 (22, 28) | 25 (22, 28) |
| | Obese, BMI ≥30 (%) | 6 (18.2) | 28 (14.7) |
| Current smoker (%) | | 2 (5.6) | 25 (12.5) |
| Cardiovascular disease (%) | | 2 (5.6) | 4 (2.0) |
| Antihypertensive medication (%) | | 7 (19.4) | 14 (7.0) |
| Kidney disease (%) | | 1 (2.8) | 1 (05) |
| Diabetes mellitus (%) | | | |
| | Yes, type 1 | 0 (0.0) | 2 (1.0) |
| | Yes, type 2 | 0 (0.0) | 5 (2.5) |
| | Yes, gestational | 0 (0.0) | 0 (0.0) |
| | Yes, other | 0 (0.0) | 0 (0.0) |
| | Yes, type not specified | 1 (2.8) | 5 (2.5) |
| Lung disease (%) | | | |
| | Yes, asthma only | 0 (0.0) | 15 (7.5) |
| | Yes, both asthma and lung disease | 0 (0.0) | 2 (1.0) |
| | Yes, lung disease only | 0 (0.0) | 1 (0.5) |
| | Yes, type not specified | 2 (5.6) | 21 (10.5) |
| Current cancer (%) | | 0 (0.0) | 1 (0.5) |
| Immunosuppressive medication[3] (%) | | 3 (8.3) | 6 (3.0) |
| Healthcare professional (%) | | | |
| | Interacts with patients | 3 (8.3) | 26 (13.0) |
| | Does not interact with patients | 2 (5.6) | 12 (6.0) |
| Months entering study (%) | | | |
| | April-May 2020 | 31 (86.1) | 164 (82.0) |
| | June-July 2020 | 3 (8.3) | 17 (8.5) |

| | | | |
|---|---|---|---|
| | August-September 2020 | 2 (5.6) | 8 (4.0) |
| | October-November 2020 | 0 (0.0) | 11 (5.5) |
| | December 2020-January 2021 | 0 (0.0) | 0 (0.0) |
| | February 2021 | 0 (0.0) | 0 (0.0) |
| Number of daily reports[2] | | 4 (2, 14) | 4 (1, 21) |
| Duration of study participation, days[2, 4] | | 8 (0, 82) | 10 (0, 82) |
| PCR test[5] (%) | | 5 (13.9) | 22 (11.0) |
| Antibody test[5] (%) | | 4 (11.1) | 13 (6.5) |

[1]Row percentage, [2]Median (first and third quartile), [3]Corticosteroids, methotrexate and/or biological agents (treatment of cancer and/or rheumatic disease), [4]From first to last daily report, [5]At any time during follow-up
BMI: Body Mass Index.

**Supplementary Table 8. Coefficients for model predicting symptomatic COVID-19 in COVID Symptom Study Sweden.**

| Variable | Coefficient |
|---|---|
| Loss of smell and/or taste | 2.42 |
| Fever | 1.04 |
| Unusual muscle pains | 0.71 |
| Persistent cough | 0.46 |
| Male sex | 0.40 |
| Headache | 0.26 |
| Skipped meals | 0.14 |
| Fatigue | 0.13 |
| Chest pain | 0.10 |
| Dizzy light headed | 0.00 |
| Hoarse voice | -0.03 |
| Delirium | -0.24 |
| Blisters on feet | -0.31 |
| Abdominal pain | -0.31 |
| Shortness of breath | -0.34 |
| Red welts on face or lips | -0.35 |
| Nausea | -0.37 |
| Sore throat | -0.55 |
| (Intercept) | -4.86 |
| Loss of smell and/or taste x Nausea | 0.33 |
| Loss of smell and/or taste x Diarrhoea | 0.30 |
| Loss of smell and/or taste x Dizzy light headed | 0.22 |
| Loss of smell and/or taste x Abdominal pain | 0.10 |
| Loss of smell and/or taste x Red welts on face or lips | -0.06 |
| Loss of smell and/or taste x Fatigue | -0.09 |
| Loss of smell and/or taste x Chest pain | -0.09 |
| Loss of smell and/or taste x Male sex | -0.14 |

| **Variable** | **Coefficient** |
|---|---|
| Loss of smell and/or taste x Headache | -0.16 |
| Loss of smell and/or taste x Hoarse voice | -0.28 |
| Loss of smell and/or taste x Eye soreness | -0.29 |
| Loss of smell and/or taste x Unusual muscle pains | -0.31 |
| Loss of smell and/or taste x Sore throat | -0.37 |
| Loss of smell and/or taste x Persistent cough | -0.44 |
| Loss of smell and/or taste x Fever | -0.75 |

**Supplementary Table 9. Questions on symptoms included in the CRUSH Covid Survey.**

<table>
<tr><th colspan="2">English translation of questions in Swedish</th></tr>
<tr><td colspan="2">Loss of smell and/or taste</td></tr>
<tr><td colspan="2">Shortness of breath</td></tr>
<tr><td colspan="2">Unusual fatigue</td></tr>
<tr><td colspan="2">Fever</td></tr>
<tr><td colspan="2">Loss of appetite</td></tr>
<tr><td colspan="2">Persistent cough</td></tr>
<tr><td colspan="2">Diarrhoea</td></tr>
<tr><td colspan="2">Unusual chest pain or tightness in chest</td></tr>
<tr><td colspan="2">Hoarse voice</td></tr>
<tr><td colspan="2">Stomach ache</td></tr>
<tr><td colspan="2">I’m feeling confused, drowsy, or disoriented</td></tr>
<tr><td colspan="2">Skin rashes</td></tr>
<tr><td colspan="2">Other (mark)</td></tr>
<tr><td></td><td>Runny nose/congested nose</td></tr>
<tr><td></td><td>Sore throat</td></tr>
<tr><td></td><td>Headache</td></tr>
<tr><td></td><td>Joint or muscle pains</td></tr>
<tr><td></td><td>Free text (add)</td></tr>
</table>

**Supplementary Table 10. Coefficients for the time-dependent model predicting symptomatic COVID-19 in COVID Symptom Study Sweden.**

| Variable | Coefficient |
|---|---|
| Loss of smell and/or taste | 2.67 |
| Fever | 1.15 |
| Unusual muscle pains | 0.85 |
| Persistent cough | 0.54 |
| Male sex | 0.47 |
| Skipped meals | 0.32 |
| Headache | 0.17 |
| Hoarse voice | 0.17 |
| Eye soreness | 0.09 |
| Fatigue | 0.06 |
| Chest pain | 0.05 |
| Dizzy light headed | 0.05 |
| Age (years) | 0.00 |
| Diarrhoea | -0.01 |
| Blisters on feet | -0.22 |
| Red welts on face or lips | -0.30 |
| Delirium | -0.32 |
| Abdominal pain | -0.34 |
| Nausea | -0.38 |
| Shortness of breath | -0.56 |
| Sore throat | -0.60 |
| (Intercept) | -4.58 |
| Time (spline 1) | -2.80 |
| Time (spline 2) | -2.93 |
| Time (spline 3) | -0.29 |
| Time (spline 4) | 0.23 |
| Time (spline 5) | -0.12 |
| Time (spline 6) | 1.10 |
| Loss of smell and/or taste x Nausea | 0.32 |

| Variable | Coefficient |
|---|---|
| Loss of smell and/or taste x Diarrhoea | 0.28 |
| Loss of smell and/or taste x Dizzy light headed | 0.24 |
| Loss of smell and/or taste x Abdominal pain | 0.23 |
| Loss of smell and/or taste x Delirium | 0.02 |
| Loss of smell and/or taste x Age (years) | 0.00 |
| Loss of smell and/or taste x Blisters on feet | -0.11 |
| Loss of smell and/or taste x Red welts on face or lips | -0.13 |
| Loss of smell and/or taste x Skipped meals | -0.14 |
| Loss of smell and/or taste x Shortness of breath | -0.14 |
| Loss of smell and/or taste x Headache | -0.16 |
| Loss of smell and/or taste x Male sex | -0.16 |
| Loss of smell and/or taste x Fatigue | -0.16 |
| Loss of smell and/or taste x Chest pain | -0.23 |
| Loss of smell and/or taste x Eye soreness | -0.27 |
| Loss of smell and/or taste x Hoarse voice | -0.29 |
| Loss of smell and/or taste x Unusual muscle pains | -0.32 |
| Loss of smell and/or taste x Sore throat | -0.35 |
| Loss of smell and/or taste x Persistent cough | -0.45 |
| Loss of smell and/or taste x Fever | -0.78 |
| Loss of smell and/or taste x Time (spline 5) | 0.44 |
| Loss of smell and/or taste x Time (spline 3) | -0.26 |
| Loss of smell and/or taste x Time (spline 2) | -0.35 |
| Loss of smell and/or taste x Time (spline 1) | -0.36 |

| Variable | Coefficient |
|---|---|
| Loss of smell and/or taste x Time (spline 6) | -0.44 |
| Loss of smell and/or taste x Time (spline 4) | -0.50 |
